# Playing with peers: Exploring peer support mechanisms of a type 2 diabetes-specific board game

**DOI:** 10.1101/2020.09.07.20177733

**Authors:** Pil Lindgreen, Vibeke Stenov, Ingrid Willaing, Henning Grubb Basballe, Lene Eide Joensen

**Author notes:** **Corresponding author** Pil Lindgreen, Diabetes Management, Steno Diabetes Center Copenhagen, Niels Steensens Vej 2, DK-2820, Gentofte, Denmark.

## Abstract

**Aim:** To explore specific mechanisms of a board game aiming to facilitate peer support among people with type 2 diabetes.

**Design:** Qualitative observational study including game tests in group-based diabetes education programs and focus groups exploring experiences among people with type 2 diabetes playing the game.

**Methods:** The game was tested with 77 people with type 2 diabetes in March-July 2019. Data from audio recordings of game tests and focus groups were analysed using interpretive description.

**Results:** Six mechanisms facilitated peer support among people with type 2 diabetes: 1) entering a safe space of normality created by emotional in-game mirroring; 2) mutual in-game acknowledgment of out-of-game efforts; 3) forming relationships through in-game humor; 4) health care professionals using game rules to support group dialogues of interest to people with type 2 diabetes; 5) being inspired by in-game exchange of tips and tricks; and 6) co-players guiding each other during the game. Peer support was inhibited by the mechanism of game rules obstructing group dialogues.

**Conclusion:** The board game effectively facilitated peer support through playfulness and humor and helped players form relationships. Additionally, the game established a framework for players to share and listen to each other’s diabetes-specific experiences, promoting a sense of normality and cohesiveness. This process depended on facilitating educators’ flexibility to balance the rules of the game with its aim of creating peer support.

**Impact:**

- The study provides detailed knowledge on specific board game mechanisms facilitating peer support among people with type 2 diabetes participating in diabetes education programs.
- The game was an effective tool to facilitate emotional and practical peer support by encouraging players to share and acknowledge each other’s diabetes experiences.
- Peer support is promoted through the game by creating a sense of normality and belonging among people with type 2 diabetes.

## 1. INTRODUCTION

In Denmark, an estimated 240,000 people are diagnosed with type 2 diabetes (T2D) (Danish Health Authority, 2017). People with diabetes have a significant risk of developing severe physiological diabetes complications and reduced psychosocial functioning (Perrin et al., 2017; World Health Organization, 2016), which may impair diabetes self-management and adversely affect outcomes (Joensen, Almdal, et al., 2016; World Health Organization, 2008). Due to treatment costs and reduced quality of life, diabetes poses a substantial challenge to health care systems (International Diabetes Federation, 2017; World Health Organization, 2016).

People diagnosed with T2D (PWT2D) in Denmark are offered community-based educational programs (“Bekendtgørelse af Sundhedsloven,” 2019; Danish Diabetes Association, 2018) that typically include 6-8 weekly group sessions. Nurses, dietitians and physiotherapists present diabetes diet, exercise and medication guidelines in a classroom setting (Danish Diabetes Association, 2018), often supplemented by group activities (e.g., cooking and exercising). The programs are ideal settings for implementing tools facilitating the peer support PWT2D need to maintain diabetes self-management (Boothroyd & Fisher, 2010; Debussche et al., 2018; Fisher, Ballesteros, et al., 2015; Funnell, 2010; Harkness et al., 2010; Simmons et al., 2015; van Dam et al., 2005). The key functions of peer support are: 1) assistance in daily disease management; 2) social and emotional support; 3) linkage to clinical care and community resources; 4) ongoing support of chronic disease management (Fisher, Ayala, et al., 2015). Peer support is associated with improved diabetes management and psychosocial and physical outcomes, including reduced stress, depression and hospitalisation (Chan et al., 2014; Heisler, 2010). Moreover, people attending diabetes-specific peer support interventions highly value exchanging experiences with their peers (Christoffersen et al., 2018; Jensen et al., 2019) and may benefit as much from providing social support as from receiving it (Heisler, 2010). In general, participants appreciate peer interactions in educational group programs (Stenov et al., 2016). However, more knowledge on effective methods and interventions to enhance peer support is needed (Joensen, Filges, et al., 2016).

### 1.1 Background

Steno Diabetes Center Copenhagen and Copenhagen Game Lab collaborated to develop a board game designed to promote peer support in Danish T2D educational group programs. It was intended to enable group participants to share diabetes experiences and provide participants and facilitating HCPs with insights into group members’ diabetes-related challenges, needs and preferences. Despite the seriousness of the topic, the rationale for creating a board game was to allow peer support to flourish in a playful manner because humour can be a powerful aid to the exchange of personal experiences (Buiting et al., 2020; Schöpf et al., 2017).

Based on the literature, we hypothesised that peer support would be facilitated by mechanisms including the related concepts of mirroring, normalization and recognisability; cohesiveness and reciprocity; humour and playfulness; and sharing tips and tricks. People often better understand and verbalize their thoughts, behaviours and emotions when they are mirrored in their peers’ responses (Gruhl et al., 2015). Mirroring that occurs among people with similar experiences and challenges is closely linked to the concept of normalization—recognising themselves in others’ experiences and realising that they are not as different as they believed (Joensen, Filges, et al., 2016). Recognisability and normalization may replace feelings of isolation with the sense of belonging and being in a “safe space” (Joensen, Filges, et al., 2016; Joensen et al., 2017). The reciprocal exchange of diabetes experiences and management tips among group members may also engender empathy and a sense of group cohesiveness (Christoffersen et al., 2018; Heisler, 2010).

## 2. THE STUDY

### 2.1 Aim

The study aim was to explore board game mechanisms facilitating or inhibiting peer support among PWT2D playing the game as part of group-based T2D educational programs.

### 2.2 Design

The study was qualitative and observational with multicentre game tests and focus group interviews with PWT2D. Data were analysed using interpretive description (Thorne, 2016).

#### 2.2.1 The board game

Each participant plays as a fictitious persona. A card lists the persona’s personal circumstances (e.g., marital and employment status) and T2D-related strengths and challenges (e.g., physically active, concerned about complications) and depicts the persona as a caricature. Players take turns guiding their personas by selecting among theme cards containing tips related to diet, exercise, medication and social activities. The better the theme card fits the needs and challenges of the persona, the more points the player receives. After guiding their fictitious personas, players take turns reflecting on their strengths and challenges with T2D related to the selected theme. This alternation between reflecting on the persona’s needs and the player’s needs occurs continuously throughout the game.

PWT2D and HCPs were involved in the process of design and testing. More information on the development, programme theory and content of the board game is available elsewhere.

### 2.3 Participants

PWT2D were eligible to participate if they were: 1) diagnosed with T2D, 2) without developmental or psychotic disorders and 3) being enrolled in a T2D education program in one of nine included municipalities. Representing three of five Danish regions, municipalities were selected based on their varying socioeconomic profiles.

### 2.4 Data collection

Field and focus group interviews conducted in March-July 2019 generated the study data.

#### 2.4.1 Field tests and observations

Seventy-seven PWT2D participating in diabetes education programs led by 17 HCPs were included. We performed 19 field tests of the board game, each lasting 1-1.5 hours and including 3-5 PWT2D and 1-2 HCPs (Table 1). The game tests were audio recorded and transcribed verbatim. Two researchers were present to guide and assist with the game as needed. While observing the tests, the researchers took field notes on important contextual factors (e.g., multiple games being played in the same room) and nonverbal communication.

**Table 1.** Overview of data collection

|  | Municipality | Participants<br>(N) |  | Focus<br>group ID | Test<br>ID |
| --- | --- | --- | --- | --- | --- |
|  |  | PWT2D | HCPs |  |  |
|  | A | 3 | 2 | 1 | 1 |
|  | B | 4<sup>a</sup> | 1<sup>b</sup> | 2 | 2 |
|  | B | 4<sup>a</sup> | 1<sup>b</sup> | 2 | 3 |
|  | B | 4<sup>a</sup> | 1<sup>c</sup> | 3 | 4 |
|  | B | 4 | 1<sup>c</sup> | 3 | 5 |
|  | C | 5 | 1 | 4 | 6 |
|  | C | 4 | 1 | 4 | 7 |
|  | D | 5<sup>a</sup> | 1 | 5 | 8 |
|  | D | 5 | 1 | 5 | 9 |
|  | E | 5 | 1 | N/A<sup>e</sup> | 10 |
|  | F | 3 | 1 | 6 | 11 |
|  | F | 4 | 1 | 6 | 12 |
|  | F | 3<sup>a</sup> | 1 | 6 | 13 |
|  | G | 3 | 1<sup>d</sup> | 7 | 14 |
|  | G | 4 | 1<sup>d</sup> | 7 | 15 |
|  | H | 4<sup>a</sup> | 1 | 8 | 16 |
|  | H | 4<sup>a</sup> | 1 | 8 | 17 |
|  | I | 4 | 1 | 9 | 18 |
|  | I | 4 | 1 | 9 | 19 |
| **Total**<br>**(N)** | **0** | **76** | **17** | **9** | **19** |

<sup>a</sup> A spouse accompanied one group participant.
<sup>b</sup> One HCP simultaneously facilitated the games of test 2 and 3.
<sup>c</sup> One HCP simultaneously facilitated the games of test 4 and 5.
<sup>d</sup> One HCP simultaneously facilitated the games of test 14 and 15.
<sup>e</sup> Focus group interview not conducted due to time restraints.
Abbreviations: HCP, healthcare professional; PWT2D, person with type 2 diabetes

#### 2.4.2 Focus group interviews

Nine focus group interviews were conducted to explore PWT2D’s experiences of the game. A semi-structured interview guide addressed overall experience and satisfaction with the game, insights gained from discussions of diabetes-related topics and perception of the group atmosphere during the game. Focus group interviews were audio recorded and transcribed verbatim.

### 2.5 Ethical considerations

The study was approved by the Data Protection Agency (project ID: VD-2018-157) and conducted according to the Helsinki Declaration (WMA General Assembly, 2018, July 9) and current Danish legislation. No ethical approval was needed (The National Committee on Health Research Ethics, 2011). Participants provided informed consent after receiving verbal and written information about the study aim and methods and their right to withdraw at any time without affecting their future treatment.

### 2.6 Data analysis

Data were organised and coded using NVivo 12®. Data analysis was an iterative four-step process (Thorne, 2016). The researchers first agreed on an initial coding structure reflecting the purpose of the game, then continuously revised it as new categories were identified and others were combined or eliminated (Suppl. material). All data coded was completed by three researchers. Second, data unrelated to the study aim (e.g., on HCPs’ experiences) were discarded (Thorne, 2016). Third, preliminary themes grounded in the remaining data were formulated through iterative assessment and discussion. Fourth, the final themes were fully developed (Thorne, 2016).

### 2.7 Validity and rigour

The validity of the findings was enhanced by data from different sources and perspectives (Sandelowski, 1995; Thorne, 2016). The study aim was investigated objectively through observations of game play and subjectively by exploring players’ experiences.

The rigour of the findings was ensured by continuous discussions of emerging findings. When disagreements arose, the raw data was reviewed to identify specific data points supporting or refuting preliminary findings (Thorne, 2016).

A semi-structured interview guide was developed based on key literature within the topics of diabetes education programs and gamification strategies and on the programme theory of the game. Different types of questions, such as probing, specifying and structuring, were included (Kvale, 2014).

## 3. FINDINGS

Participants were an average of 64 years old (range, 42-89 years) and 53% (n=39) were male. Of all participants, 44% (n=31) had one or more chronic diseases in addition to T2D, 85% (n=60) took oral hypoglycemic agents and 18% (n=10) injected insulin. More details on participant characteristics are available elsewhere.

Specific game elements (i.e., personas and theme cards) introduced different mechanisms that facilitated or inhibited peer support. Two types of peer support were identified: emotional peer support for diabetes-specific psychosocial challenges (e.g., worries about future complications) and practical peer support related to the instrumental requirements of diabetes self-management (e.g., medication adherence) and the game itself (e.g., guiding co-players about the rules). Emotional and practical peer support were facilitated by different mechanisms but inhibited by a single mechanism (Figure 1).

**Figure 1:**
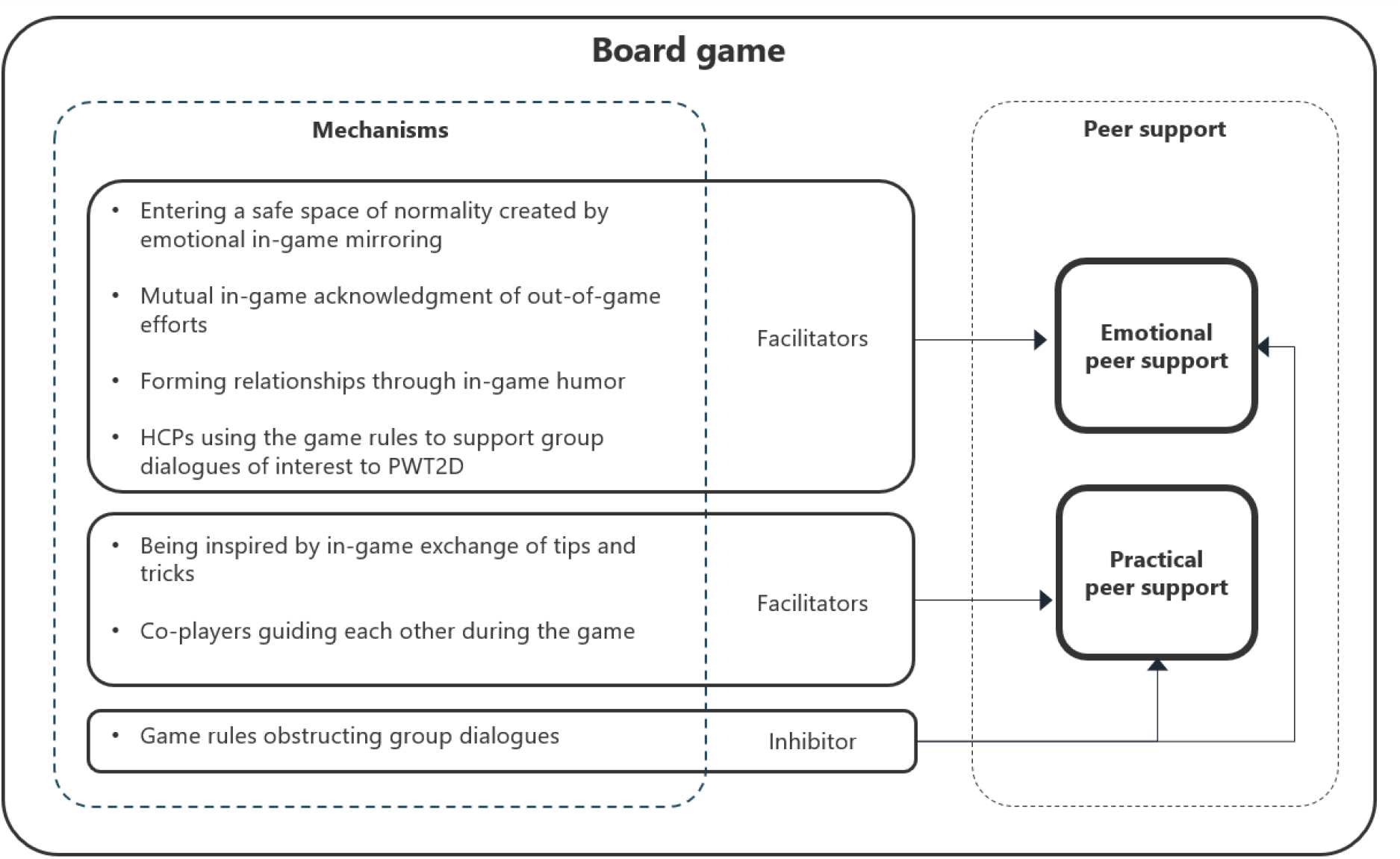
Illustration of the identified mechanisms of the board game that facilitated or inhibited emotional and practical peer support.

### 3.1 Mechanisms facilitating emotional peer support

Emotional peer support was demonstrated when participants shared and respectfully responded to diabetes-specific emotions elicited by game elements.

#### 3.1.1. Entering a safe space of normality created by emotional in-game mirroring

##### 3.1.1.1 In-game mirroring

The game rules directed players to compare themselves to their personas. This mirroring encouraged the players to reflect on, verbalise and openly share their emotional responses to and challenges with diabetes. A participant articulated: “[…] it can be good to say it out loud. No matter what you’re up against” (Focus group [FG] 9). Participants highlighted similarities and differences between themselves and their personas:

> “[…] Jens [persona] cannot feel his diabetes and doesn’t talk about his illness, but he is worried about complications. He is afraid of not being able to live as he is used to [...] I don’t talk much about the illness either [...] but I’m quite worried about complications, so I’m trying to learn more about it. So I can postpone them a bit.” (Test [T] 9)

Emotional peer support was also observed when co-players mirrored themselves in each other’s narratives as illustrated in field notes: “[…] They listen to and acknowledge each other” and “The participants look at each other and nod when others are talking. They seem interested in what the others are saying” (T5). Similarly, one participant said: “[…] what I like about this [game] is that you gain insights into your own experiences and become able to share them […] And listen to what the others are saying [...] we were very open about everything” (FG2).

##### 3.1.1.2 Feeling normal

Mutual mirroring enabled participants to share diabetes-related emotions and engendered a sense of normality by acknowledging the fact that challenging diabetes-related emotions are common. As one participant said:

> “[…] we have different lives and different approaches, but you can still recognize yourself in a lot of what is being said. And it’s nice to know that you’re not the only one with these thoughts […] it’s all related to the disease somehow.” (FG9)

Another participant in the same group said, “You almost become normal.” The sense of normality was also observed: “[...] phrases like ‘I feel the same way as you’ and ‘Me too’ are being used” (T4). The process of exchanging diabetes experiences also engendered a sense of group cohesiveness in participants: “[…] it [playing the game] creates a sort of community [...] we share the same kind of things. It gives you a sense of belonging to the group” (FG9).

##### 3.1.1.3 Entering a safe space

Group cohesiveness was enhanced by game rules directing players to take turns. In addition to equalising the amount of time spent on each player, taking turns ensured that all players contributed their experiences in addition to listening to those of others. This exchange created a safe space: “[…] some of the more vulnerable things that are sometimes difficult to talk about are definitely brought up this way. And then you get a little closer in the group somehow, you feel safer around each other” (FG9). Participants felt increasingly comfortable sharing experiences that they would not otherwise speak about. “You start talking about some things, you wouldn’t normally talk about,” a participant said, to which another responded, “Yes, and it’s not something you’d talk about with a co-worker. That goes for several of these [theme] cards” (FG2).

#### 3.1.2 Mutual in-game acknowledgment of out-of-game efforts

When discussing their choice of theme cards for their personas, participants often shared their diabetes experiences, including self-management efforts outside the game (i.e., in “real life”), which their co-players then acknowledged. One example of emotional peer support occurred when a participant said: “[…] I put the green one [game piece] on ‘taking the stairs’ [...] I had one of those pedometers, where it exceeded 15,000,” to which a co-player responded, “Oh boy, that’s a lot! That’s bloody well done” (T3). Similarly, when discussing diet, one participant described planning healthy meals that could be prepared in the oven. While the food cooked, she went for a walk as part of her daily exercise routine. A co-player acknowledged her planning skills: “I find it admirable that you’re thinking that far ahead [...] I’m impressed by that” [...] I think you deserve a big pat on the back for that” (T18).

Participants acknowledged each other’s diabetes management strategies, regardless of how they were actually implemented. This approval replaced guilt, often associated with less-than-perfect compliance with official diabetes guidelines, with acceptance from the group and contributed to establishing a safe space. A participant said: “[…] for those experiencing it [guilt] I think they become able to let go of some of their guilt. They get a pat on the shoulder and acceptance” (FG9). For example, when discussing a theme card on exercise, one participant said that she skipped her walks in bad weather but took other measures to stay active: “I don’t go for walks when it’s rainy and windy [laughs] But I’m doing well with the fitness thing and actually showing up there [...] twice a week.” A co-player replied, ‘That’s good, and a great insight [...] Instead of walking around constantly feeling guilty, then you’re telling yourself that’s just the way it is” (T17).

#### 3.1.3 Forming relationships through in-game humour

In focus group interviews, several participants described the game as “fun”. Researchers also noted participants’ obvious enjoyment: ‘They laugh during the game, especially when discussing the theme cards that are sometimes very familiar” (T1). Participants generally joked throughout the game, especially about their personas. For instance, a participant chose a theme card advising his persona to join a cooking class to learn diabetes-friendly recipes while socializing; a co-player responded wittily: “It’s a boring cooking class he’s in if he doesn’t get to have any red wine” (T6). Humour also arose when participants played personas very different from themselves. For example, a male participant proclaimed his lack of interest in gardening when comparing himself to his female persona, who enjoyed garden work. Later, when selecting a theme card on exercise for his persona, the participant ironically noted: “I clean often and spend time gardening. It’s great exercise.” The group responded with laughter (T13).

The use of humour contributed to a light-hearted atmosphere and promoted emotional peer support as participants bonded, sometimes by highlighting shared preferences. Two participants joked about their preference for driving over walking: “The car needs exercise, too,” one said, to which the other replied, “And you get exercise when moving your foot.” They both laughed, and the first participant added, “It [the foot] also needs exercise” (T2). Forming relationships through humour added to the sense of group cohesiveness: “[…] the playfulness and the lack of restraint [...] playing a game does something when it comes to getting to know one another,” said one participant (FG9). Similarly, a participant said, ”[…] it [the board game] was inspiring [I had some] nice input from the -”. Another participant interrupted with: “The members.” The group laughed, and the first participant continued, “Yes, the members. That’s a little bit cool. You’re safe in an environment, where everyone knows what it’s about” (FG8). Humour contributed to the sense of a safe space, as exemplified by this exchange (FG8) that was followed by good-natured group laughter.

Participant 1: [...] I’m usually not open about my diabetes [...] I don’t think I’ve told anyone besides you guys. I think of it as something private.

Participant 2: We won’t gossip about it.

Participant 1 (sarcastically): Now, are you sure about that?

#### 3.1.3 HCPs using game rules to support group dialogues of interest to PWT2D

HCPs also facilitated emotional peer support. By listening attentively to participants’ diabetes-specific needs and preferences, they ensured that group dialogues were relevant to everyone. If a dialogue took an unproductive turn, HCPs redirected participants’ attention to something new. This was legitimized by referring to the game rules, which preserved a friendly atmosphere. For instance, when a participant delivered a monologue that could have disengaged co-players from the game, the HCP interrupted her to steer the game back on track:

> “But I think you’re going to talk about all of that soon. Because you’re doing a lot. And that’s amazing. But right now, I think you [the group] have to proceed with it [the game] and then later, you [the group] will tell us about your experiences” (T8).

Another way for HCPs to keep dialogues interesting to participants was to highlighting similarities between their experiences. This occurred during a discussion of a theme card that described the option to eat the same food as everyone else at social events to avoid standing out.

> Participant: “If you’re going out [...] It’s easier not making a fuss about it and simply eating what’s being served [...] than getting all that attention. I can’t do it.”
>
> HCP: “You mentioned the negative attention you get when eating something else. Does the rest of you experience that, too? I mean, how do you manage it? How about you [name]? you’ve had it [TD2] for many years, right?” (T10).

Similarly, another HCP emphasized similarities between participants’ experiences after a discussion of a theme card about physical activity: “So, there are some things you have in common. Having some arrangements to exercise with people is helpful, because having an appointment and spending time with others motivate you” (T13).

However, HCPs sometimes inhibited emotional peer support by dominating the game and replying to participants’ questions or remarks instead of letting co-players respond. One participant considered a theme card for her persona with the option to spend time with grandchildren to momentarily forget about T2D, who was described as highly concerned about future diabetes complications (T13):

Participant: Well, she’s an elderly lady, and some exercise would do her good.

HCP: She’s worried.

Participant: Yes, she’s worried about the illness. And, I don’t know if she has grandchildren –

HCP: She has four grandchildren”

Participant: Four grandchildren. And when she spends time with them, she’s not thinking about her disease.

HCP: So she gets a break from her disease?

Participant: Yes.

### 3.2 Mechanisms facilitating practical peer support

Practical peer support was provided when participants exchanged diabetes management tips and tricks as inspiration and guidance.

#### 3.2.1 Being inspired by in-game exchange of tips and tricks

Sharing specific diabetes management strategies provided practical peer support. Typically, it occurred when participants discussed their rationale for choosing a specific theme card. For instance, a participant commented on a theme card suggesting replacing sugary candy with a few pieces of dark chocolate:

> “[…] The one [theme card] about dark chocolate. It’s the chips [for me]. But I found a solution to that, because I found out that I can make some dip using sour cream [...] then I can eat carrots and cauliflower […] instead of chips” (T2).

By relating the theme card to a different personal challenge, the participant shared a diabetes management tip in enough detail that others could adopt it. In another example (T6), a participant (1) chose a theme card on a strategy he had used of regularly eating fish with low-fat dressing because he valued healthy food that was tasty. Two co-players (Participants 2 and 3) drew a parallel between this behaviour and making sugar-free desserts:

Participant 2: I like something sweet, right. Participant 1: Me, too. Participant 2: And the Danish Diabetes Association has some good desserts. You just have to buy this strange kind of sugar [...] artificial sweetener. Participant 1: Oh, I didn’t know about that. Participant 3: Yes, you can simply add a bit of sweetener. Participant 1: Well, I see, but I don’t have any. I have to go buy it now. Participant 3: Yes, you’ll have to go buy some. Participant 1: Yes, I see. There are so many options.

By sharing this tip, the two participants inspired their co-player to adopt another diet-related behaviour. Similarly, a participant told the group how she struggled to eat enough vegetables because her husband did not like them. A co-player offered a potential solution: “Have you tried hiding them [the vegetables] in lasagna and stuffing?” (T18).

The exchange of specific strategies and tricks inspired participants. One said: “What I liked about this [game] was how we asked each other ‘why are you doing that, or could you do this, and how would you do at your house’ [...] you got a lot of ideas” (FG1). These exchanges highlighted similarities between participants’ diabetes management strategies, increasing group cohesiveness. As one said: “It’s easier to talk to others who are in the same situation [...] you’re not alone in the world” (FG7).

#### 3.2.2 Co-players guiding each other during the game

Some participants actively guided co-players on the game rules (e.g., helping them understand the point system). In addition to helping game progress, game-specific guidance contributed to a friendly atmosphere and the creation of a safe space, as illustrated in field notes: “The participant reading aloud the rules is very pedagogical. She explains the rules, using examples. She is especially helpful to one co-player who finds the game a bit challenging” and “The participants are friendly and helpful towards each other” (T5). For example, one participant (P5) was unsure how to select a theme card:

P5: But there are no more [theme cards] that fit him [persona].
P6: What kind of person is he?
P5: He is busy, and -
P6: Do you remember what his description said?
P5: Well, he’s busy and he’s [pauses] Shouldn’t I be talking about myself?
P6: No. You have to tell us what [theme] card you chose for him and [...] why [...] Do you want me to go first? Is that better?
P5: Yes, that’s better.
P6: If not, then say so.
P5: I will. It’s a bit difficult for me.

### 3.3 Mechanism inhibiting emotional and practical peer support

#### 3.3.1 Game rules obstructing group dialogues

Emotional and practical peer support were inhibited by the same mechanism. Peer support was inhibited when following the rules and completing the game were prioritized at the expense of meaningful group dialogues. This typically occurred when time was short; participants limited discussions to ensure they would finish the game. A participant said: “[…] you feel like you have to get through it [the game]. Then, there isn’t a lot of room for individual talking” (FG8). Participants preferred having enough time to let group dialogues unfold rather than worrying about completing the game: “[...] you’d have a better outcome, if you had a bit more time” (FG8). Furthermore, when advancing the game took priority, some participants felt they took too much time sharing their experiences. For example, a participant described worrying about long-term complications because she had trouble adhering to diabetes-friendly foods. While acknowledging the relevance of her concern, the HCP also pointed out the need to continue the game to make sure everyone had a say. “Yes, of course. It’s not all about me”, the participant replied (T18).

Difficult or confusing rules also interfered with group dialogues by disengaging participants from the game: “It was difficult getting started. The rules were difficult”, one participant stated (FG5). Several participants didn’t know when to play their personas and when to be themselves.

This interfered with meaningful dialogues as the conversation shifted from diabetes-related issues to the game rules, illustrated in a discussion about what to eat when attending social events (T6):

P6: Are you talking about your persona or yourself?
P7: No, about me.
P6: But we’re not supposed to talk about ourselves, are we?
HCP2: Afterwards. Right now, we’re talking about the cards for [the persona].
P7: I did both already.
P6: You did?

## 4. DISCUSSION

We explored specific mechanisms of a board game that facilitated or inhibited peer support among PWT2D. The game facilitated emotional and practical peer support by promoting a sense of cohesiveness, normality, and acceptance among participants. However, emotional and practical peer support was inhibited when game rules were prioritized over group dialogues.

Mechanisms facilitating emotional peer support differed from those facilitating practical peer support. However, both types of peer support were inhibited by the same mechanism, indicating their interrelated nature. This is consistent with previous research demonstrating that exchanging practical diabetes management tips adds to the foundation of trust needed for emotional, or psychosocial, peer support (Fisher, Ballesteros, et al., 2015; Joensen et al., 2017). Similarly, interventions facilitating emotional peer support can engender a sense of belonging to the group that may encourage participants to share instrumental advice (Kowitt et al., 2015). Emotional peer support may gradually emerge during a peer support intervention (Fisher, Ballesteros, et al., 2015). Consequently, group dialogues may be limited if game play occurs in initial group sessions, but early game play may also encourage participants to share diabetes-specific experiences and emotions with peers early on.

Entering a safe space of normality created by emotional in-game mirroring and being inspired by in-game exchange of tips and tricks respectively promoted emotional and practical peer support. Similar findings have been reported by Joensen et al., who found that people with type 1 diabetes experienced diabetes-specific social capital when exchanging diabetes-related experiences and tips, which reduced loneliness and induced a feeling of being “normal” (Joensen, Filges, et al., 2016; Joensen et al., 2017). Co-players guiding each other on how to play the game (e.g. by explaining the game rules) while playing it facilitated practical peer support along with a sense of cohesiveness. This is similar to the processes described by Hessler (2010) by which reciprocal guidance from “insiders” (i.e. peers participating in the same group or activity) enhances a sense of belonging among peers (Heisler, 2010). Thus, the board game is a highly relevant way of promoting group cohesiveness and structured peer support.

Mutual in-game acknowledgment of out-of-game efforts facilitated emotional peer support, in part by reducing feelings of diabetes-specific guilt (Hilliard et al., 2015; Joensen et al., 2017; Sebire et al., 2018). Similarly, conversations with HCPs that incorporate dialogue tools supporting articulation of the difficulty of daily diabetes management can also reduce diabetes-specific guilt (Jensen et al., 2019). Discussions with like-minded peers, however, may more effectively reduce guilt because of shared experiences and understanding (Heisler, 2010).

Emotional peer support was also facilitated by forming relationships through in-game humour facilitated by the game (e.g., by its illustrations). Similarly, other studies have found that humour plays a significant role in the formation of peer support in interventions for various medical conditions. Among people with chronic pain, collectively generated humour on relatable topics (e.g., side effects of analgesics) made participants feel included as group “members” (Finlay et al., 2018). Rabin (2018) found that young adult cancer survivors frequently used humour when sharing their cancer history with peers to keep the topic as “light” as possible. The humorous aspect of peer support can also exist outside of face-to-face group meetings. A review of online communities for people with diabetes concluded that peers frequently shared humorous diabetes-related content (e.g., personal anecdotes or comics) as a way of coping with the seriousness of living with diabetes (Hilliard et al., 2015). Among women with gynaecological cancer, “black” or “gallows” humour during telephone-based peer support contributed significantly to peer bonding (Pistrang et al., 2012). HCPs can thus consider various online, telephonic, and face-to-face ways of facilitating peer support in their practices. In addition to humour shared with peers, people with diabetes may benefit from humour shared with HCPs. Studies have shown that humour can positively affect patient-provider interactions by reducing the power asymmetry and promoting a more relaxed atmosphere (Schopf et al., 2017). However, Schöpf and colleagues (2017) pointed out that the use of humour was most successful when initiated by patients rather than the provider, who should simply mirror it. Similarly, women with gynaecological cancer legitimized morbid humour when it was delivered by peers, not HCPs (Pistrang et al., 2012). In all, humour is a highly effective tool to facilitate peer support by building and strengthening peer relationships, thus resulting in a sense of belonging. Moreover, when initiated by patients, light humour may benefit patient-provider relationships (Schöpf et al., 2017).

HCPs using the game rules to support group dialogues of interest to PWT2D facilitated emotional peer support, whereas game rules obstructing group dialogues inhibited both emotional and practical peer support. Similarly, a study of various dialogue tools in T2D education programs found that the tools primarily promoted participant dialogues, but could also obstruct them (Jensen et al., 2019). This was the case when tools were too complicated or participants could not relate to the content (Jensen et al., 2019). Similarly, we found that game rules mostly served the purpose of encouraging group dialogues but occasionally did the opposite when players did not understand them. Thus, to promote peer dialogue through board games, careful instructions need to be readily available for facilitating HCPs.

### 4.1 Limitations

Several limitations deserve mention. Video recordings would have allowed us to completely document nonverbal communication. However, we recorded field notes while observing the game tests (Thorne, 2016). The transferability of our findings is limited by the fact that our study was carried out in Denmark, where T2D educational programs and healthcare are free. Moreover, the specific cultural setting, for instance regarding the acceptance of humour and open discussion of emotions, may impact the outcomes of playing the game, which should be considered when transferring the board game to other settings. A study strength is the analysis by three researchers, who repeatedly discussed the coding structures and findings to maintain rigour (Thorne, 2016). Additionally, data from observations and focus group interviews explored game mechanisms from different perspectives. We also collected data from several settings, which increased representativeness and potential transferability of the findings (Thorne, 2016).

## 5. CONCLUSION

Our study provides novel insights into the mechanisms of board-game play that facilitated or inhibited peer support among PWT2D participating in T2D educational programs. The game promoted emotional and practical peer support in a playful and humorous way, encouraging participants to engage in structured group dialogues. The exchange of diabetes-specific experiences, needs, challenges and management tips induced a sense of normalization, acceptance, and belonging among PWT2D. The facilitation of peer support depended on HCPs’ abilities to balance encouraging group dialogues with adhering to the game structure. HCPs need training and instructions on how to achieve the needed flexibility.

## Data Availability

The data are not publicly available due to privacy or ethical restrictions.

## Acknowledgements

We want to express our gratitude to the people with type 2 diabetes participating in the study and the health care professionals present during the game tests for letting us gain access to their diabetes education program sessions, enabling us to study the mechanisms of the board game in practice; we will do our best to ensure that our findings will benefit people with type 2 diabetes in general. Furthermore, we would like to thank Copenhagen Game Lab for their extensive collaboration throughout the process of designing, testing, and evaluating the board game. Finally, we want to acknowledge research assistants, Thit Hjortskov Jensen and Maria Friis Børsting, for their assistance in collecting, transcribing, and analyzing the data.

## Funding Statement

Financial support to conduct the study was provided by Steno Diabetes Center Copenhagen. No external funding was received.

## Conflicts of interest statement

The authors have no conflicts of interest to report.

## Notes

### Competing Interest Statement

The authors have declared no competing interest.

### Clinical Trial

The study is registerd at Center for Open Science with the following identifier: DOI 10.17605/OSF.IO/ZHJ5E

### Author Declarations

The study was approved by the Data Protection Agency (project ID: VD-2018-157) and conducted according to the Helsinki Declaration (WMA General Assembly, 2018, July 9) and current Danish legislation.

